# Confirmed COVID-19 cases per economic activity during Autumn wave in Belgium

**DOI:** 10.1101/2021.05.31.21256946

**Authors:** Johan Verbeeck, Godelieve Vandersmissen, Jannes Peeters, Sofieke Klamer, Sharon Hancart, Tinne Lernout, Mathias Dewatripont, Lode Godderis, Geert Molenberghs

## Abstract

**Objective:** To assess the COVID-19 incidence per economic activity during the Autumn wave 2020 in Belgium.

**Methods:** The 14-day incidence of confirmed COVID–19 cases per NACE–BEL code is described in the periods immediately preceding the Belgian more strict measures of October 19, 2020, and is evaluated longitudinally by a Gaussian–Gaussian modelling two–stage approach. Additionally, the number of high-risk contacts in working segments and regions is described.

**Results:** The peak of COVID–19 14–day incidence in most NACE–BEL sectors is reached in the period October 20-November 2, 2020 and was considerably higher than average in human health activities, residential care activities, fitness facilities, human resource provision, hairdressing and other beauty treatment and some public service activities. Human health activities, residential care activities, food and beverage service activities, hotels, arts, food retail activities, and human resources provision have high pre-lockdown incidences. The frequency of index cases that report more than two high risk contacts is increasing over time in all sectors.

**Conclusion:** Despite the restrictive protocols present in many sectors before the Autumn wave, employees in activities where close contact with others is high, show increased risk of COVID–19 infection. Especially sports activities are among the highest risk activities. Finally, the increasing amount of high-risk contacts by COVID–19 confirmed cases is compatible with the decreasing motivation over time to adhere to the measures.

**Key Messages:** 

**What is already known about this subject?:** Certain occupational sectors, such as human health and care, food and beverage, cultural and sport activities, have been related to a high risk of SARS-CoV-2 infection at the workplace.

**What are the new findings?:** COVID-19 confirmed cases of employees are linked with the main economic activity of their employer. The effect of opening of sectors, potentially under restrictive protocols, is evaluated. Despite the restrictive protocols present in many sectors, employees in activities where close and/or prolonged contact with others is high exhibit increased risk of COVID–19 infection, even higher than the high-risk sector of human health and care. Full restriction of these sectors decreases adequately the COVID-19 incidences, even in those sectors with physical contacts that remain open, for example human health, care and food shops. Finally, the increasing amount of high-risk contacts by COVID–19 confirmed cases might be related to decreasing motivation over time to adhere to the measures.

**How might this impact on policy or clinical practice in the foreseeable future?:** These insights offer guidance to policy makers on which economic activity to restrict or subject to stricter protocols to better control the COVID-19 pandemic whilst keeping the work floor as safe as possible.

## Introduction

Since the SARS-CoV-2 virus was identified in Wuhan (Hubei, China) in December 2019, the virus spread globally, causing the largest pandemic in a century. Managing this pandemic was a challenge for many countries. Soon it was clear that the virus was airborne and spread through close human contact. In Italy it was estimated that, up to May 2020, COVID-19 was probably contracted at work in 30% of cases at working age.[2] Many countries implemented interventions to limit physical contact in private life and at work. As little was known on the role of various actors and activities on the spread of SARS-CoV-2 at the beginning of the first wave, a general lockdown to manage spread and prevent health care system collapse was introduced by many governments. Despite adequately controlling viral spread, a full lockdown leads in time to serious economic and well-being issues; hence, more targeted tools are necessary, such as the restriction and re-opening of economic activities. Therefore, it is essential to gather knowledge on the dynamics of the virus and the risk of contracting COVID-19 in different economic activities.

Economic activities where social distancing is challenging, have been related to clusters of COVID-19 cases. In Japan, 61 clusters of COVID-19 were tracked to health care (30%) and other care (16%) facilities, cultural activities (11%), gyms (8%), ceremonies (3%) and transport (2%).[3] COVID-19 outbreaks have been documented in economic activities of Manufacturing, Agriculture/Forestry/Fishing/Hunting, and Transportation/Warehousing in the US[4] and Canada,[5] while several sources report COVID-19 outbreaks in poultry, meat and food processing companies [6] and residential care facilities.[7, 8] Employees in bars and restaurants have been shown to have increased COVID-19 risk [1, 9] or were involved in clusters of COVID-19 [10]. This was confirmed by a study of the European Centre for Disease Control (ECDC) that examined the number of clusters per sector during the first wave in 15 European countries and the United Kingdom.[11] In Norway, the odds of COVID-19 was 1.1–3 times higher during the first wave in nurses, doctors, dentists, physiotherapists, bus, tram, and taxi drivers relative to the general population at working age.[1] During the second wave, however, the odds did not increase for some contact professions suggesting that taking appropriate measures at work can contain the spread of the virus at the workplace.

Despite the observed association between activities and COVID-19, it does not provide information on the effect of opening or closing sectors on the spread of SARS-CoV-2. In some US states, closing and re-opening of bars, restaurant and schools and wearing masks were found to have a significant effect on the spread of the virus, hospitalisations and deaths. [12, 13] A study combining mobility data with confirmed COVID-19 cases examined the effect of re-opening single economic activities on viral spread[14]: restaurants, gyms, hotels, cafes, and religious organizations were identified to produce the largest increase in infections when re-opened under pre-pandemic conditions. Therefore, many policy makers have decided to allow re-opening of these locations only with reduced visitors and visitor time to control SARS-CoV-2 spread. To our knowledge, only one study investigated different opening strategies. In selected municipalities in Norway, bartenders and waiters had similar rates of COVID-19 in areas with full and partial bans on serving.[9]

In Belgium, a general, national strict lockdown was installed on 18 March 2020, closing schools and suspending all cultural, leisure, and non-essential activities.[15] These extreme measures proved successful in decreasing the daily new infections and COVID-19 hospitalisations to a level where the government felt confident to gradually alleviate the lockdown measures. While most activities could resume over Summer 2020 with strict protocols, including restrictions on capacity, dwelling time, and social contacts, in September 2020 these restrictions were relaxed despite indication of increased circulation of SARS-CoV-2.[15] Since October 2020, progressively more restrictive measures were implemented to contain the second COVID-19 wave, ultimately leading to a closure of bars and restaurants on 19 October, and further tightening on 2 November (closure of social and some economic activities; no school closure nor movement restriction).[15] Arguably due to the consistency of measures and restrictions on end of year festivities, Belgium managed to avoid further flare-ups until March 2021.[16].

By linking COVID-19 confirmed cases of employees with the NACE-BEL code [17] of the main economic activity of their employer, the COVID-19 incidence is examined over Autumn and Winter 2020. A cross-sectional analysis of the COVID-19 14-day incidence immediately prior to 19 October 2020 is contrasted with a longitudinal analysis of the incidence over the entire Autumn, to examine the effectiveness of the soft lockdown. Finally, contact tracing of confirmed COVID-19 cases gives insights into the high-risk contacts in specific work segments and regions.

## Methods

### Data

The Belgian institute for health, Sciensano registers daily all confirmed COVID-19 cases, [18] and forwards them to the National Social Security Office (NSSO), who in turns links these to the Dimona database of active employees. The Dimona database covers most of the employees (∼ 4.5 million), such as employees in private and public sectors, interim employment and student job, but includes neither self-employed nor foreign workers that are not subjected to the Belgian social security scheme.

The data are aggregated at NSSO to weekly incidences (number of cases per 100,000) by NACE-BEL code. NACE-BEL classifies workplaces into 21 main economic sectors (level 1) and then further into ever finer subcategories (levels 2 – 5), with 943 subcategories at level 5.[17] Although some companies may be active in more than one sector, only the main NACE-BEL code is assigned. This limitation is particularly important for education, because a majority of schools provide both primary and secondary education, while all employees are categorized as secondary school personnel.

As the code is given at company level, no distinction is made between activity within the company (e.g., administrative work in metal industry). No information is available on exact employment location (omitting information on telework or temporary unemployment).

Finally, the actual source of infection, particularly workplace or elsewhere, is unavailable. Hence, the data are useful to compare the incidence evolution with overall trends in the working population and in the general population.

Data on workplace high-risk contacts of confirmed COVID 19 cases are available from the IDEWE contact tracing database. IDEWE is one of the largest occupational health services in Belgium and responsible for the well-being of approximately 800,000 employees from 33,000 companies or institutions, covering more than 20% of Belgian workers. IDEWE is active in all economic sectors, with a predominance in healthcare. More than 20% of all employees, under medical surveillance of IDEWE, are working in the healthcare sector.

Since 29 October 2020, the COVID-Contact Tracing application, developed by IDEWE for its employers, registers in a standardized manner information on COVID-19 incidences and on high- and low-risk contacts of index cases. Most of the index cases are employees, but also seasonal workers, interns, pupils and external people working at companies’ and institutions’ premises that tested positive are contacted. Contact tracing of high- and low-risk contacts, defined according to Sciensano guidelines [19], within the company is performed. Measures are taken for within-company high-risk contacts (testing, 7 or 10 days quarantine). Contacts exterior to the company are identified and followed-up through the regular contact tracing. Employers are grouped by customer segment in one of 9 regional offices, named after the city where they are located. Most Belgian provinces have one regional office, except Antwerp that is served by the regions Antwerpen, Mechelen, and Turnhout; and Namur that serves all of Wallonia. IDEWE distinguishes between ten customer segments based on NACE codes, but an exact link with the NSSO codes is not fully possible. Some larger IDEWE companies have organized contact tracing via their internal prevention service, which is not included in this analysis, potentially leading to underestimation of index cases. For some segments this underestimation might be more important.

### Statistical analyses

The NSSO weekly incidences from 8 September 2020 to 25 January 2021 are mapped to 14-day incidences by joining two consecutive weeks. Adjacent 14-day incidences share an overlapping week. Details on the calculation of the 95% confidence interval for the incidence is available in online supplementary annex A.

For the 5 NACE-BEL levels, the highest incidences in the two 14-day periods before the measures of 19 October 2020 (29 September–12 Oct 2020; 6–19 October 2020) are presented, together with the 14-day incidence over all work sectors (∼ 4.5 million individuals) and in the general population (∼ 11.5 million individuals).

The longitudinal profile of the 14-day incidences is modelled by fitting so-called Gaussian-Gaussian functions, in a two-step approach (online supplementary annex B).

Precision in small NACE-BEL sectors is low. Hence, for levels 1, 2, and 3, only sectors with a minimum of 10,000 employees are analyzed, for levels 4 and 5, the minimum is 3000 and 1500, respectively.

For the index cases that were reported via IDEWE tracing between 29 October 2020 and 18 February 2021, the mean number of high-risk contacts and the four-weekly percentage of index cases with two or more high-risk contacts are described per work segment and per region. Under-reporting may arise because the tracing application reports zero high-risk contacts for an index case by default, which might be incorrect for an index that is non-contactable or refuses to respond. Over-reporting might occur in the education segment. The contact tracing for schools is performed by Student Guidance Centres (SGC), who forward the contract tracing of pupils to IDEWE if employees might be involved as high- or low-risk contact. The SGC tracing is centre dependent and often only index cases with high-risk contacts are forwarded to IDEWE.

## Results

### Pre-peak period

At NACE-BEL level 1, among sectors with a minimum of 10,000 employees, Arts, entertainment and Recreation; Accommodation and food service activities; Human health and social work activities; Public administration and defence, compulsory social security; and Education show a 14-day incidence above the NACE-BEL average in both periods before 19 October 2020 (Figure 1 and online Supplementary Table 1).

**Figure 1:**
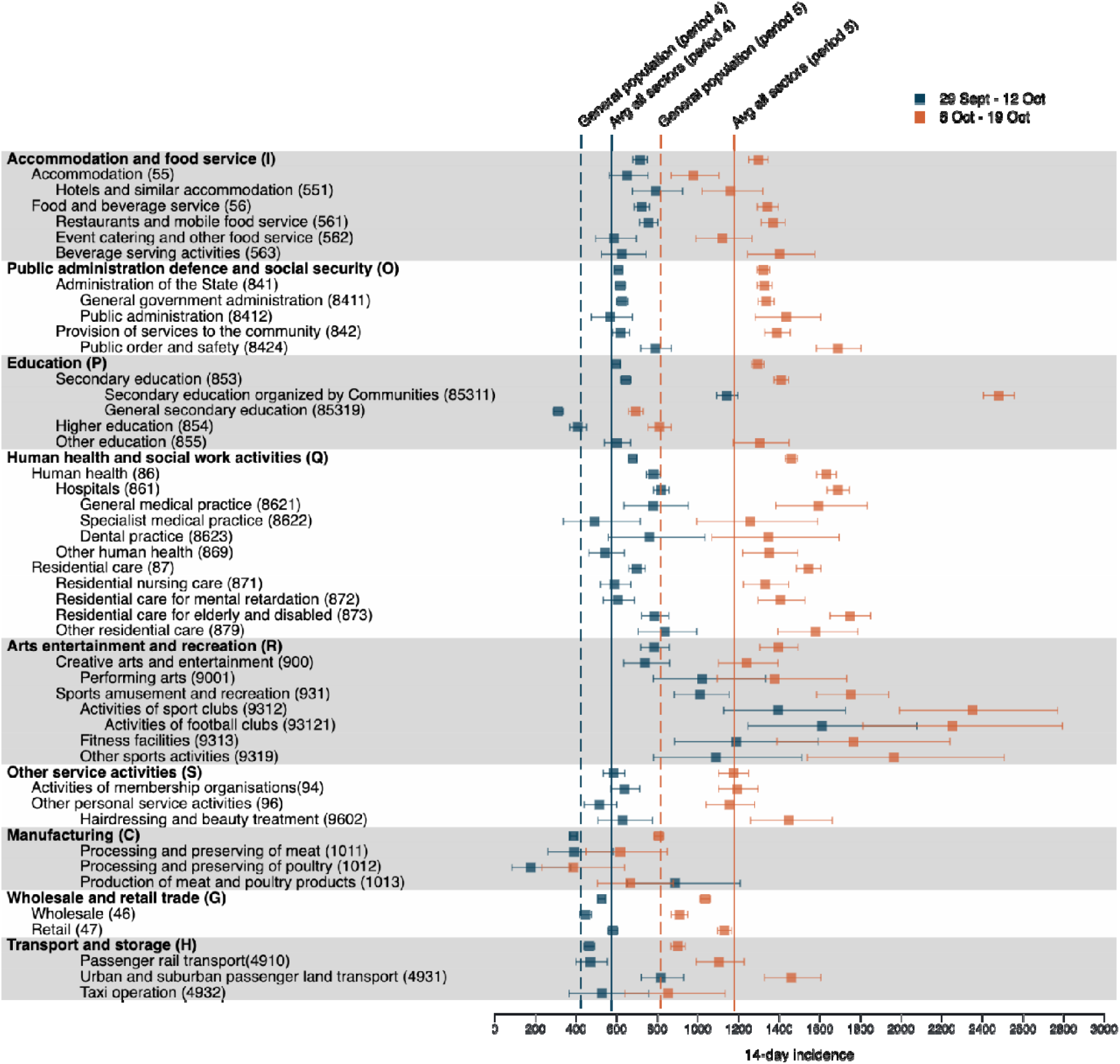
14-Day incidence of COVID-19 in selected sectors in Period 29 September – 12 October and 6 – 19 October

From levels 3 – 5 it follows that within the Arts, entertainment and Recreation sector, mainly Sports activities have high incidence before 19 October 2020 (Figure 1 and online Supplementary Tables 2 and 3). Within sports, high incidences come from activities of sport clubs (especially football clubs,), fitness activities, and other sport activities (Figure 1 and online Supplementary Table 4 and 5). As the human health and social work sector is subject to frequent close contacts, it is no surprise that its incidence is among the highest among the lower level sectors (Figure 1 and online Supplementary data). The sports activity sector has higher incidence still. Also, education organized by the regional authorities (sector 85311) has a higher incidence than the care sector, unlike general education (sector 85319) and other levels of education (sector 854, 855) (Figure 1 and online Supplementary data).

The Accommodation and food service incidence is comparable to that of health and care, and is similar between hotels, restaurants, and bars between 29 September–12 October, while hotels are doing slightly better between 6–19 October (Figure 1 and online Supplementary data). Within Public administration, defence and social security, the incidence in governmental and general administration services is above the all-sector average, while the Federal and local police in the public order and safety sector have the highest incidences at level 1 (Figure 1 and online Supplementary file). Within the Other service activities, the incidence of the non-medical contact professions stands out with incidences similar to the health and care sector (Figure 1 and online Supplementary file).

The spread of COVID-19 in meat and poultry processing sectors (sector 1011, 1012, 1013) seems well controlled in the periods immediately prior to measures, with incidence slightly lower than in the general population (Figure 1 and online Supplementary file). Incidences in Wholesale and retail trade are well controlled before 19 October 2020, although some sectors show an increased incidence (sectors 4729, 4631, 4764, 4773) (Figure 1 and online Supplementary file). In Transportation and storage, incidence in the urban and suburban passenger surface transportation is higher than average.

All NACE-BEL sectors with their corresponding 14-day incidence and confidence interval can be found in the online Supplementary file. Various rankings or aggregates of sectors can be easily constructed from this file.

The pre-peak period can further be studied via the longitudinal profile, which is represented by the pre-peak plateau parameter δ_*s*1_ in the Gaussian-Gaussian model. Although the plateau parameter at level 1 indicates no significant differences in incidences before the peak, the incidences in Sports activities, more specifically Activities of sport clubs and Residential care for elderly and disabled are significantly elevated (Figure 2). At level 4, additionally Child-day care, Organisation of conventions and trade shows, and some Wholesale and retail trade sectors have an increased incidence before the peak (Figure 2).

**Figure 2:**
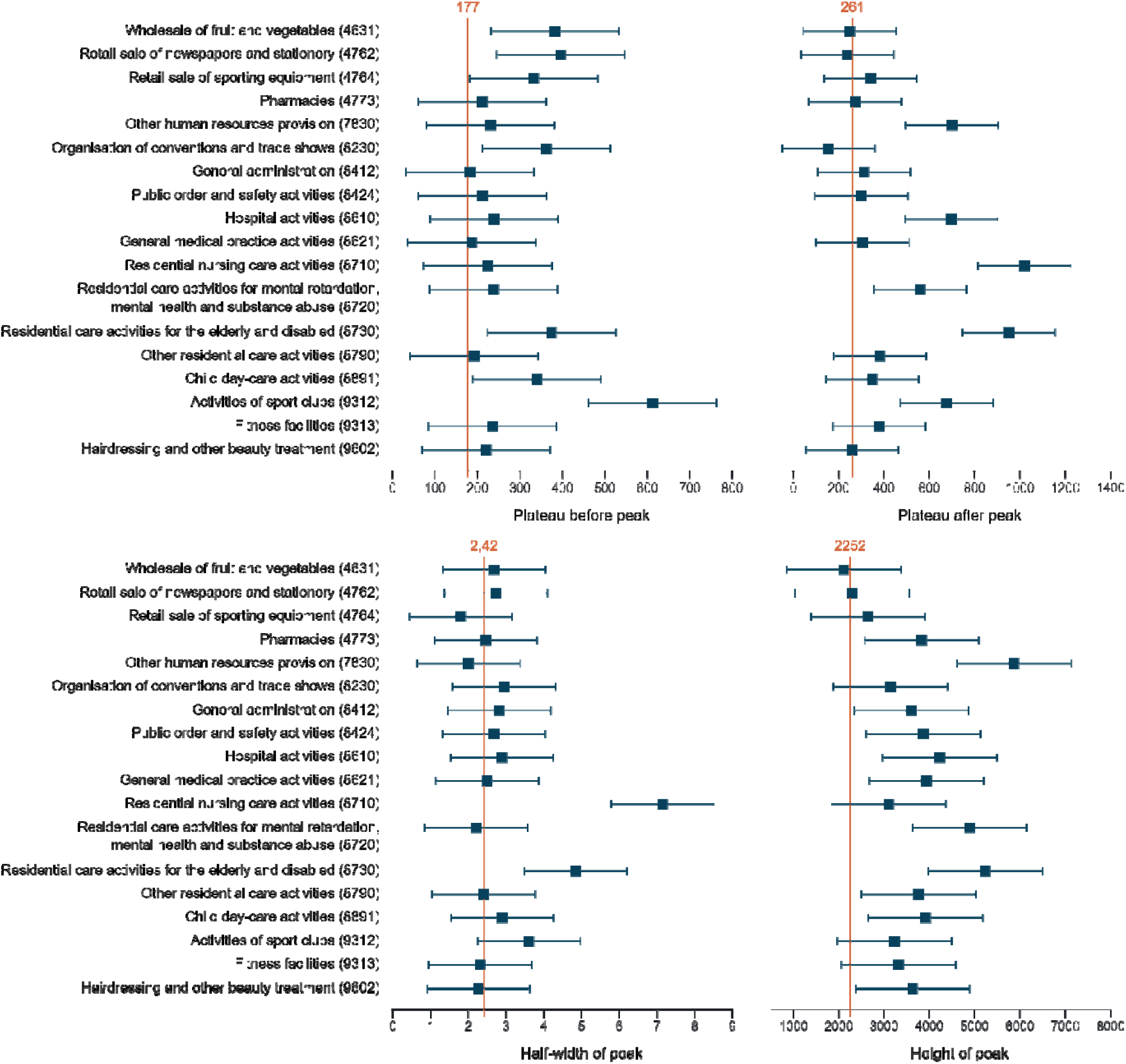
Forest plots of characteristics of the longitudinal profile of selected sectors. The plateau before and after the peak are related to the 14-day incidence. The height of the peak is the 14-day incidence at the highest moment in the curve and the half-width of the peak is the number of weeks it takes for the curve to reduc**e** the 14-day incidence by a half.

### Peak of the Autumn wave

Both the cross-sectional and longitudinal analyses show that for the majority of sectors the Autumn wave reaches the incidence peak in the period of 20 October–2 November 2020 (Figure 3 and online Supplementary file).

**Figure 3:**
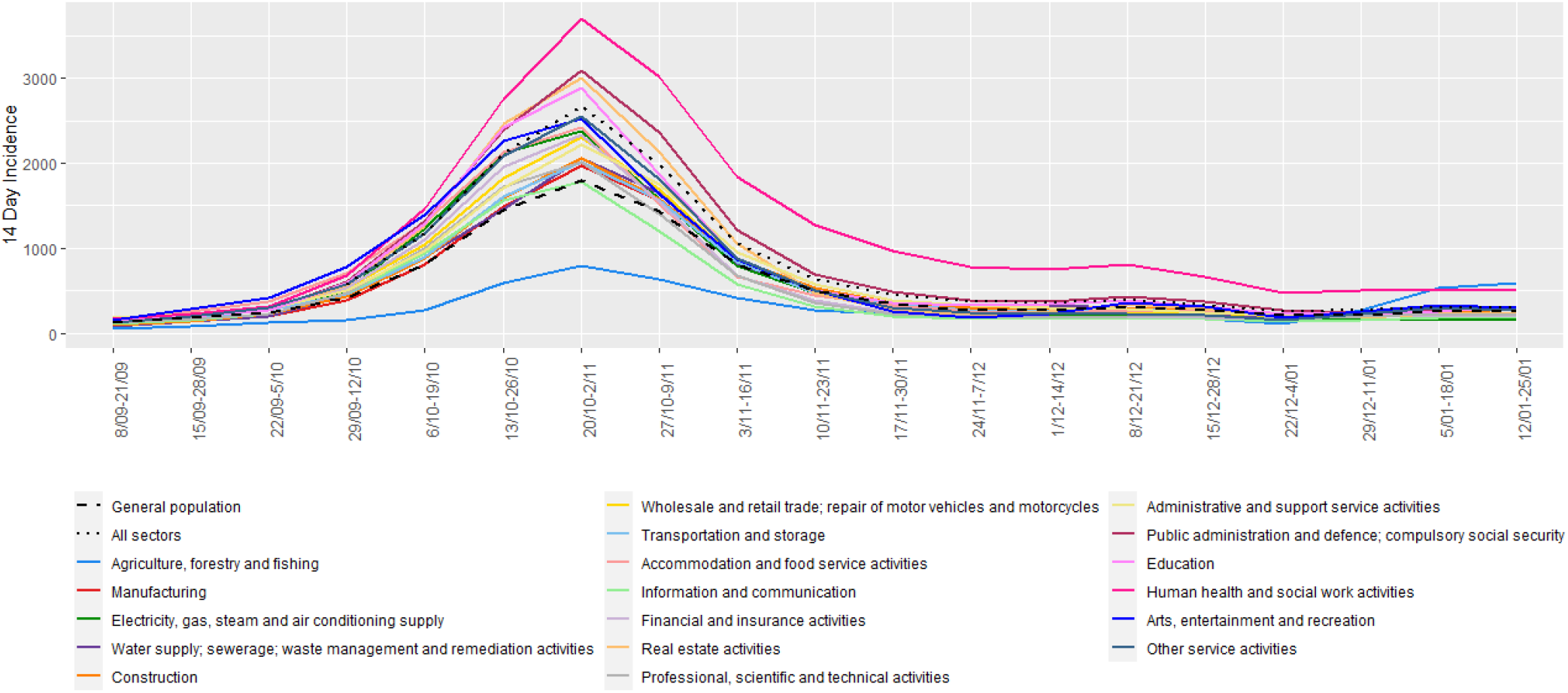
14-Day incidence of COVID-19 in sectors with minimum 10,000 employees at Level 1

The longitudinal analysis shows that the peak is significantly higher for Human health and social work activities, both for the Human health activities, and the Residential care activities (Figures 2 and 3). Within Human health activities, both Hospitals and General medical practice have a higher incidence, while for Residential care activities, most sectors show an extreme peak (sectors 872, 873, 879). At level 4, additionally Fitness centre activities, non-medical contact professions, General administration, Federal and local police, Pharmacies, and Other human resource provision have a significantly elevated peak incidence (Figure 2).

### Post-peak period

In the longitudinal analyses two parameters inform us about the incidences after the peak. The plateau after the peak, captures the incidence to which the sector decreased, while the half-width, quantifies the time in weeks for the incidences to decrease.

Human health and social work activities, both for the Human health activities, and the Residential care activities, have a significantly higher post-peak incidence level, while the latter sector also has a larger half-width (Figures 2 and 3). Within Human health activities, Hospitals have a higher post-peak incidence, while for the Residential care activities, most sectors show both an increased incidence after the peak (sector 871, 872, 873), and a longer half-width (sector 871, 873). At Level4, Activities of sports clubs, and Other human resource provision (sector 7830) had a significantly elevated post-peak incidence (Figure 2). For most sectors the post-peak plateau is higher than the pre-peak one (Figure 4).

**Figure 4:**
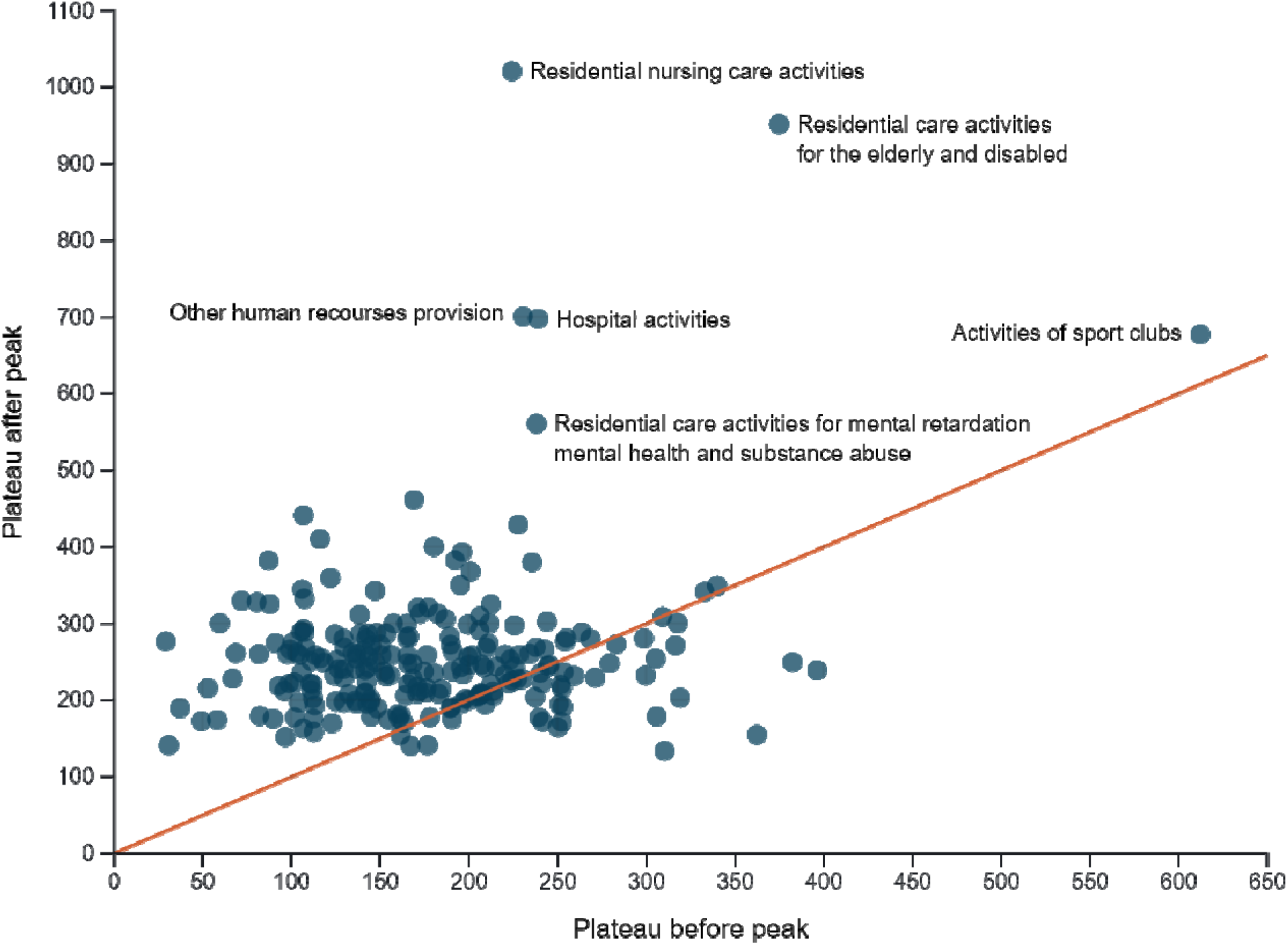
Comparison of the plateau before and after the peak in sectors at Level 4.

### Contact tracing

From 29 October 2020 until 18 February 2021, high-risk contacts of 6,016 index cases were recorded. The number of high-risk contacts per index case varied from 0 to 56, with more than 99% having less than 10 high-risk contacts. For about 5952 indexes, customer segment and region were known. The frequency of index cases that reported two or more high-risk contacts increased over time in most segments and all regions (Figure 5). Index cases in the segments Emergency services, Education, Government, and Public transport reported a higher mean number of high-risk contacts over this period, compared to other segments, while Health care and Accommodation & Food trade and industry index cases reported relatively low values (online Supplementary Figure 6). The mean number of high-risk contacts reported was higher in Hasselt compared to other regions (online Supplementary Figure 7).

**Figure 5:**
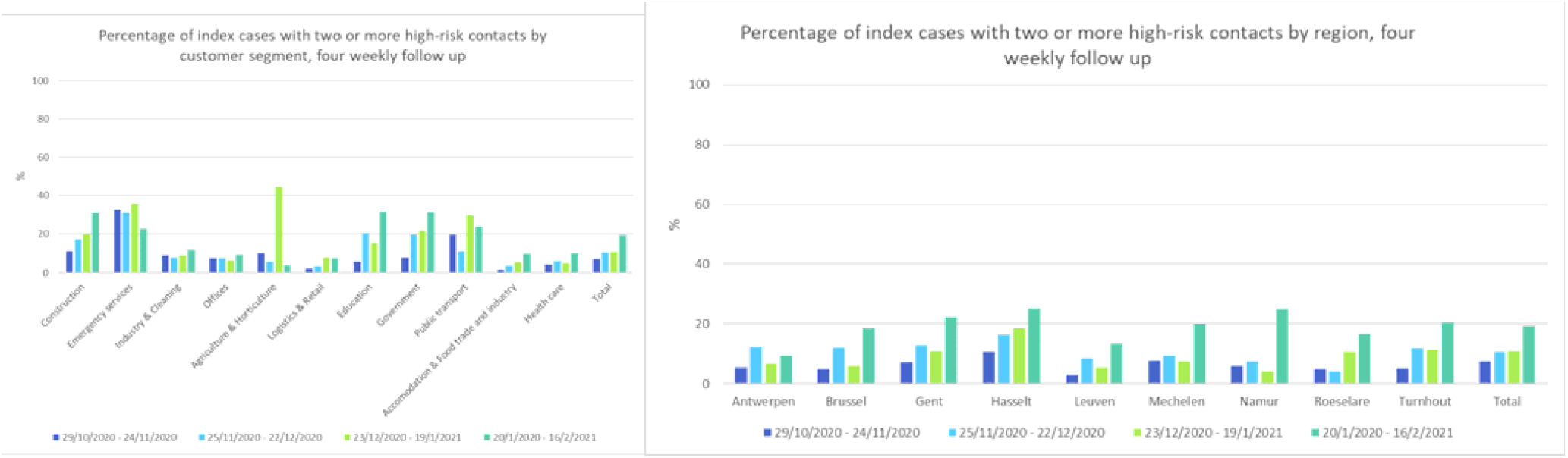
Four-weekly percentage of index cases with two or more high-risk contacts by segments under surveillance (left) and by geographical region (right)

The number of index cases for the segments ‘construction’, ‘emergency services’ and ‘agriculture’ and for the region ‘Namur’ was very low during some time periods, leading to imprecise estimates.

## Discussion

We studied the 14-day incidence of COVID-19 per economic activity in Belgium, cross-sectionally, during the uptick of the Autumn wave and longitudinally during the entire wave. Measures prior 19 October 2020 [15] are described in online supplementary annex C.

With the September protocols in place, incidence was increased in: sports activities; hotels, bars and restaurants; arts; public transport; certain types of stores; child day-care; public law enforcement; and education during the build-up of the Autumn wave. In many of these sectors, incidence exceeded that of naturally high risk-sectors such as human health activities and residential care activities. Sports activities, strongly driven by football club activities, had the highest incidence at almost all levels before 19 October 2020.

An relevant metric is a sector’s peak incidence. Apart from human health activities and residential care activities, also other human resource provision, fitness facilities, hairdressing and beauty treatment and some public services are high-peak activities. Before the peak, again human health activities, residential care activities, but also sports activities had a significantly elevated incidence.

Economic activities with increased incidence are mostly sectors with the professional need for close proximity to other people. Based on the risk inflation factor suggested by Jobs At Risk Index (JARI) [21], in occupations that bring employees into close contact with others and/or with infections, such as in health care activities, residential care, prison and undertakers, a higher COVID-19 incidence is expected. However, in Belgium sectors that are labelled by JARI as occupations with only close proximity and no regular contact with disease (education, law enforcement, fitness, beauty, retail, musicians/actors, restaurants and bars, and transport) have an equally high or further elevated incidence. Since index cases in the health care segment report relative low amount of high-risk contacts, this suggests that health care employees are effective in avoiding high-risk contacts and/or health care infection protective protocols are efficient. The restrictive rules before the second wave may have been insufficient for the close proximity occupations. Arguably, employees’ behaviour in these sectors could be more risky (on the work floor and/or beyond), as evidences by increased reporting of high-risk contacts by Public transport for example. Additionally, restrictive protocols may be sufficient during periods of low-level virus circulation but progressively less with increasing incidence.

Our results contrast with the findings of no increased incidence in sports activities of the SafeActive survey on self-reported COVID-19 in a sample of fitness and exercise facilities [22] and that of no difference in incidence in a sample of occupations in a UK survey [23]. Our findings agree with the analysis of mobility data, identifying gyms, bars and restaurants as a high-risk location of infection, [14] and the reports of clusters of COVID-19 and outbreaks [1, 3, 4, 5, 6, 7, 8, 9, 10]. Various high-incidence sectors are mentioned as potential location of infection by index patients during contact tracing. Places mentioned most as activity or event visited two weeks before infection are restaurants and bars, sports activities, public activities, wellness and hairdressers and fitness facilities (Flemish contact tracing). While no formal proof for the place of infection, the increased incidence in these sectors is striking.

On 19 October 2020 more stringent measures were issued in Belgium to control the emerging Autumn COVID-19 wave [15] (online supplementary Annex D). On 2 November 2020, non-essential shops, non-medical contact professions, bars, and restaurants professions were closed. Hotels remained open. The effect is these measures is seen in the timing of peak incidence, which for most sectors is in the 2-week period past 19 October 2020, for the general population, sectors with restricted activity, and sectors that remained active, such as human health activities, residential care activities, and essential shops. Despite their forefront position [24] employees in the food industry seem adequately protected and well informed on protective measures in Belgium, as incidences in food retail decreased to the all-sector average after the peak; the number of high-risk contacts reported by Accommodation & food trade and industry is low.

The effect of the measures is also seen in width and height of post-peak incidence. Unsurprisingly, the peak width is significantly broader for human health activities and residential care activities. For most activities, post-peak incidence is higher than pre-peak, reflecting controlled but increased SARS-CoV-2 circulation. Human health activities, residential care activities, activities of sports clubs and other human resource provision have a significantly higher post-peak incidence. Although incidence in sports activities is largely influenced by virtually unrestricted activities of professional football clubs, also fitness facilities, other sports activities, and activities of leagues and sports federations have an increased peak and/or pre-lockdown incidence.

The success of the measures to curb the second wave notwithstanding, the increasing number of index cases over time reporting 2 or more high-risk contacts potentially demonstrates the decreasing motivation to adhere to these measures. Evidently, increasing high-risk behaviour by a part of the general population may result in delays towards relieving non-pharmaceutical interventions by the decision makers.

Further insights on the COVID-19 incidence per economic activity should be gained from including information on self-employed workers.

## Strengths

The data analyzed here includes all confirmed COVID-19 cases among employees, interim employment, and job students, across all economic activities; it is thus more complete than information based on a random sample on a restricted set of occupations or a self-completed survey in a sample.[23] Besides the cross-sectional description of incidences, several aspects of the COVID-19 wave are compared via a longitudinal Gaussian-Gaussian model.

## Limitations

The absence of information on COVID-19 incidence in self-employed workers is a limitation. As the proportion of self-employed workers per NACE-BEL sector is variable, this might have variable impact. The data depends on the COVID-19 testing strategy, which has changed on 20 October 2020. To safeguard testing laboratory capacity, testing of asymptomatic individuals following a high-risk contact was suspended until 23 November 2020. However, this likely impacts most sectors equally. NACE-BEL codes are assigned only to the main activity of a company and no inference can be made regarding the location of infection (workplace or elsewhere) nor the location of employment (work, telework, temporarily unemployed). The results, however, do reflect the behaviour and potential risk of spreading COVID-19 by employees in a sector.

## Conclusion

Despite the limitations of the data, our results give clear insights in the incidence and the effect of restrictive protocols on COVID-19 incidence per NACE-BEL sector. These insights offer guidance to policy makers on which economic activity to restrict or relieve to control the COVID-19 pandemic and keep the work floor as safe as possible.

## Supporting information

Supplementary annex

## Data Availability

Cross-sectional data are included in the article or uploaded as supplementary information. Longitudinal data are available upon reasonable request.

## Contributor ship

Conceptualization by GM, JV, MD and LG. JV and GV performed the statistical analyses. JP provided the figures. SF, SH and TL prepared the data. JV drafted the paper, which was commented and agreed upon by all co-authors.

## Ethics Approval Statement

Not applicable.

