## Supplementary annex for "Confirmed COVID-19 cases per economic activity during Autumn wave in Belgium"

Supplementary Material 1

**Annex A**

**The calculation of the 95% confidence interval for the 14-day incidence**

The 95% confidence interval (CI) for the 14-day incidence is calculated on a logit transformation scale of the proportion, after which it is back-transformed to the original scale. Let *Y_st_* be the random variable representing *number of COVID-19 confirmed cases* in the 14-day period *t* for sector *s*, *y_st_* its realized value, and *p_st_* the *proportion of COVID-19 infected employees*, then *I_st_* = *p_st_ ×* 100*,*000 is the *14-day incidence*. The 95% CI of the logit transformation of the proportion

$$\hat{\lambda}_{st}=\log\left( \frac{p_{st}}{1-p_{st}} \right),$$

with variance

$\hat{\sigma}_{st}^{2}=\frac{1}{y_{st}(1-p_{st})}$_is_ $\hat{\lambda}_{st}\pm1.96 \sqrt{\hat{\sigma}_{st}^{2}}$.

By back-transforming this CI, the 95% CI of the 14-day incidence is

$$\frac{e^{\hat{\lambda}_{st}\pm1.96 \sqrt{\hat{\sigma}_{st}^{2}}}}{1+ e^{\hat{\lambda}_{st}\pm1.96 \sqrt{\hat{\sigma}_{st}^{2}}}}$$

**Annex B**

**Modelling of the 14-day incidences by fitting so-called Gaussian-Gaussian functions, in a two-step approach.**

The Gaussian-Gaussian function is an asymmetric bell-shaped curve, with different scale parameters on the left and right side of the common peak. In the first step, parameter estimators of the parameter vector $\theta_{s}=(\delta_{s1}, \delta_{s2}, \alpha_{s1}, \alpha_{s2}, \sigma_{s1}, \sigma_{s2}, \sigma_{s}^{2}, \nu_{s})$are obtained per NACE-BEL sector *s* by maximizing the Gaussian likelihood:

$$L_{s}\left( \theta_{s} | I_{s1}, \ldots, I_{sT} \right)=\prod_{t=1}^{T} \frac{1}{\sigma_{s}\sqrt{2\pi}}e^{-\frac{\left[ I_{st}-\mu_{st} \right]^{2}}{2\sigma_{s}^{2}}}$$

but with a non-linear mean function that is itself (asymmetrically) bell shaped:

$$\mu_{st}= \left\{ \begin{aligned} \delta_{s1}+\alpha_{s1}e^{-\frac{\left( t-\nu_{s} \right)^{2}}{\sigma_{s1}^{2}}} if t< \nu_{s}, \\ \delta_{s2}+\alpha_{s2}e^{-\frac{\left( t-\nu_{s} \right)^{2}}{\sigma_{s2}^{2}}} if t\geq\nu_{s}, \end{aligned} \right.$$

and $\nu_{s}$ the time at the peak in sector *s*, $\delta_{s1}+\alpha_{s1}\equiv\delta_{s2}+\alpha_{s2}$ the height of the peak, $\delta_{s1}$ and $\delta_{s2}$ the height of the incidence in the plateau phase before and after the peak, respectively, and $\sqrt{2ln(2)}\sigma_{s1}$and $\sqrt{2ln(2)}\sigma_{s2}$the half-width of the peak before and after the peak. For each NACE-BEL code the fully parameterized Gaussian- Gaussian model is fitted. When there is evidence of overparametrization, parameters left and right of the peak are set equal in steps until convergence. The order in which the parameters are set equal, whenever needed, are the $\alpha_{sj}$ , the $\sigma_{sj}$ , and the $\delta_{sj}$ (*j* = 1*,* 2), respectively, until a symmetric Gaussian curve is obtained.

In a second step, the variance-covariance matrix of the parameter estimators is used to construct the 95% confidence intervals (CI) for the parameter estimators. Extreme parameter values are identified as those where the CI is excluding the mean of the parameter estimates.

Note that, in a fully hierarchical analysis, the vector $\theta_{s}$ would be decomposed in a fixed and random part, and a non-linear mixed-effects model constructed. While this would take the correlation between observations in the same sectors into account in the most principled fashion, the two-stage approach is considered an adequate and computationally feasible approximation.[20]

**Annex C**

**Restrictions in Belgium in September 2020**

On 1 September 2020, the following restrictions applied in Belgium:

• Wearing a mask is mandatory in public places;

• Teleworking (working from home) is recommended;

• Social bubble of close contacts consists of 5 people per month;

Shopping is possible as a couple, with no time limit, but with a maximum capacity applicable to the shop;

• Restaurants and bars are open, with minimal distance between tables of 1.5 meters and sanitary protocols; • Sport facilities are open with restrictions on capacity and with sanitary protocols;

• Primary schools are open without restrictions. Secondary schools are open with face mask mandate for pupils and teachers. Higher education is open with restrictions on capacity per lecture hall and mask mandate for students and teachers;

• Public events are allowed with a maximum of 200 people indoor and 400 outdoor.

On 23 September 2020, these rules were adapted to allow shopping without maximum capacity and replaced the social bubble of 5 contacts to unlimited contacts, but a maximum of 10 adults per social gathering.

**Annex D**

**Restrictions in Belgium from 19 October.**

• Close contacts are limited to 1 person;

• Private and public gatherings are limited to 4 people;

• Teleworking is mandatory for all occupations where this is possible;

• Bars and restaurants are closed;

• A curfew is installed between midnight (earlier in some regions) and 5am, with a ban on alcohol sales from 8pm onward;

• Indoor activities can continue under existing protocols;

• Audience for sports events are halved from 400 to 200 spectators.

These rules were further restricted in two steps. On 23 October 2020, audiences were banned for sports events, class occupancy rate reduced in higher education, the capacity of indoor cultural events reduced and amusement parks and zoological gardens closed.
