## Supplementary Tables and Figures for "Confirmed COVID-19 cases per economic activity during Autumn wave in Belgium"

### Supplementary materials 2

Table 1: 14-Day incidence of COVID-19 in sectors with minimum 10,000 employees at Level 1 period 4 and 5

| Period 4: 29 September–12 October 2020 | | |  |
| --- | --- | --- | --- |
| DESCRIPTION | NACE Code | Employees | Incidence (95%CI) |
| Arts, entertainment and recreation | R | 60382 | 785(718;859) |
| Accommodation and food service activities | I | 219468 | 714(680;750) |
| Human health and  social work activities | Q | 588348 | 678(657;699) |
| Public administration and defence; compulsory social security | O | 542270 | 608(588;629) |
| Education | P | 531993 | 597(577;618) |
| Other service activities | S | 81336 | 584(534;639) |
| **All sectors** |  | **4390750** | **573(566;580)** |
| Real estate activities | L | 24599 | 561(475;662) |
| Financial and insurance activities | K | 125758 | 528(489;570) |
| Wholesale and retail trade;  repair of motor vehicles and motorcycles | G | 634221 | 526(508;544) |
| Administrative and support service activities | N | 350493 | 507(484;531) |
| Electricity, gas, steam and air conditioning supply | D | 21032 | 504(417;609) |
| Information and communication | J | 124635 | 479(442;519) |
| Professional, scientific and technical activities | M | 199790 | 476(447;507) |
| Transportation and storage | H | 272414 | 464(439;490) |
| Water supply; sewerage;  waste management and remediation activities | E | 35281 | 445(381;520) |
| Construction | F | 226328 | 433(407;461) |
| **General population** |  |  | **423** |
| Manufacturing | C | 542636 | 387(371;404) |
| Agriculture, forestry and fishing | A | 40373 | 161(126;205) |
| Period 5: 6–19 October 2020 | |  |  |
| DESCRIPTION | NACE Code | Employees | Incidence (95%CI) |
| Human health and  social work activities | Q | 587069 | 1358(1329;1388) |
| Arts, entertainment and recreation | R | 62169 | 1266(1181;1357) |
| Public administration and defence; compulsory social security | O | 556718 | 1206(1177;1235) |
| Real estate activities | L | 24568 | 1205(1076;1349) |
| Accommodation and food service activities | I | 223310 | 1202(1158;1248) |
| Education | P | 542963 | 1193(1164;1222) |
| **All sectors** |  | **4390832** | **1178(1168;1188)** |
| Electricity, gas, steam and air conditioning supply | D | 21026 | 1122(989;1274) |
| Other service activities | S | 81781 | 1103(1034;1177) |
| Financial and insurance activities | K | 124922 | 1058(1003;1117) |
| Wholesale and retail trade;  repair of motor vehicles and motorcycles | G | 638003 | 965(942;990) |
| Professional, scientific and technical activities | M | 198992 | 926(885;969) |
| Administrative and support service activities | N | 354068 | 913(882;944) |
| Information and communication | J | 125087 | 909(858;963) |
| Water supply; sewerage;  waste management and remediation activities | E | 35462 | 863(772;965) |
| Transportation and storage | H | 274308 | 847(814;882) |
| Construction | F | 226163 | 821(784;859) |
| **General population** |  |  | **816** |
| Manufacturing | C | 547403 | 741(719;764) |
| Agriculture, forestry and fishing | A | 40099 | 217(176;268) |

Table 2: 14-Day incidence of COVID-19 of 10 sectors with the highest incidence at Level 2 period 4 and 5

| Period 4: 29 September–12 October 2020 | |  |  |
| --- | --- | --- | --- |
| DESCRIPTION | NACE Code | Employees | Incidence (95%CI) |
| Sports activities, amusement and recreation activities | 93 | 26911 | 955(846;1078) |
| Human health activities | 86 | 261154 | 780(747;814) |
| Creative, arts and entertainment activities | 90 | 21922 | 739(634;861) |
| Food and beverage service activities | 56 | 191978 | 723(686;762) |
| Residential care activities | 87 | 162464 | 698(659;740) |
| Accommodation | 55 | 27454 | 652(563;754) |
| Activities of membership organisations | 94 | 47887 | 639(571;714) |
| Activities auxiliary to financial services and insurance activities | 66 | 30351 | 626(543;721) |
| Security and investigation activities | 80 | 20096 | 622(522;741) |
| Office administrative, office support and other business support activities | 82 | 36731 | 618(543;704) |
| **All sectors** |  | **4390750** | **573(566;580)** |
| **General population** |  |  | **423** |
| Period 5: 6–19 October 2020 | |  |  |
| DESCRIPTION NACE Code | | Employees | Incidence (95%CI) |
| Sports activities, amusement and recreation activities 93 | | 26687 | 1660(1513;1820) |
| Human health activities 86 | | 261680 | 1631(1583;1680) |
| Residential care activities 87 | | 162476 | 1543(1484;1604) |
| Food and beverage service activities 56 | | 189940 | 1342(1291;1395) |
| Public administration and defence; compulsory social security 84 | | 542998 | 1321(1291;1352) |
| Security and investigation activities 80 | | 20015 | 1304(1156;1471) |
| Education 85 | | 534158 | 1294(1264;1325) |
| Activities auxiliary to financial services  66 and insurance activities | | 30322 | 1273(1153;1406) |
| Real estate activities 68 | | 24582 | 1257(1125;1404) |
| Office administrative, office support and  82 other business support activities | | 37037 | 1242(1134;1360) |
| **All sectors** | | **4390832** | **1178(1168;1188)** |
| **General population** | |  | **816** |

Table 3: 14-Day incidence of COVID-19 in sectors with the highest incidence at Level 3 Period 4 and 5

| Period 4: 29 September–12 October 2020 | | |  |
| --- | --- | --- | --- |
| DESCRIPTION | NACE Code | Employees | Incidence (95%CI) |
| Sports activities | 931 | 21131 | 1008(882;1152) |
| Other residential care activities | 879 | 15412 | 837(705;994) |
| Hospital activities | 861 | 210745 | 819(781;858) |
| Hotels and similar accommodation | 551 | 20076 | 792(678;925) |
| Residential care activities for the elderly and disabled | 873 | 66667 | 786(722;856) |
| Restaurants and mobile food service activities | 561 | 149471 | 756(713;801) |
| Activities of call centres | 822 | 10133 | 750(599;938) |
| Creative, arts and entertainment activities | 900 | 21922 | 739(634;861) |
| Other passenger land transport | 493 | 40751 | 719(641;806) |
| Medical and dental practice activities | 862 | 22695 | 705(604;823) |
| **All sectors** |  | **4390750** | **573(566;580)** |
| **General population** |  |  | **423** |
| Period 5: 6–19 October 2020 | |  |  |
| DESCRIPTION | NACE Code | Employees | Incidence (95%CI) |
| Sports activities | 931 | 21062 | 1752(1583;1938) |
| Residential care activities for the elderly and disabled | 873 | 66609 | 1746(1649;1848) |
| Hospital activities | 861 | 210960 | 1688(1634;1744) |
| Other residential care activities | 879 | 15472 | 1577(1392;1786) |
| Medical and dental practice activities | 862 | 22765 | 1454(1306;1618) |
| Office administrative and support activities | 821 | 11777 | 1435(1235;1666) |
| Secondary education | 853 | 404968 | 1409(1373;1446) |
| Residential care activities for mental retardation, mental health and substance abuse | 872 | 39972 | 1406(1295;1526) |
| Beverage serving activities | 563 | 19500 | 1400(1244;1575) |
| Activities of call centres | 822 | 10367 | 1389(1181;1633) |
| **All sectors** |  | **4390832** | **1178(1168;1188)** |
| **General population** |  |  | **816** |

Table 4: 14-Day incidence of COVID-19 in sectors with the highest incidence at Level 4 Period 4

| Period 4: 29 September–12 October 2020 | | |  |
| --- | --- | --- | --- |
| DESCRIPTION | NACE code | Employees | Incidence (95%CI) |
| Activities of sports clubs | 9312 | 5954 | 1394(1126;1725) |
| Other human resources provision | 7830 | 4320 | 1250(959;1629) |
| Fitness facilities | 9313 | 3707 | 1187(884;1591) |
| Other retail sale of food in specialised stores | 4729 | 3312 | 1117(810;1538) |
| Other sports activities | 9319 | 3220 | 1087(781;1510) |
| Performing arts | 9001 | 5196 | 1020(780;1333) |
| Other amusement and recreation activities | 9329 | 3398 | 883(618;1260) |
| Manufacture of air and spacecraft and related machinery | 3030 | 5257 | 875(656;1166) |
| Other residential care activities | 8790 | 15412 | 837(705;994) |
| Service activities incidental to air transportation | 5223 | 5968 | 821(621;1085) |
| **All sectors** |  | **4390750** | **573(566;580)** |
| **General population** |  |  | **423** |
| Period 5: 6–19 October 2020 | |  |  |
| DESCRIPTION NACE code | | Employees | Incidence (95%CI) |
| Other human resources provision 7830 | | 4326 | 2381(1967;2880) |
| Activities of sports clubs 9312 | | 5875 | 2349(1991;2769) |
| Other sports activities 9319 | | 3208 | 1964(1537;2506) |
| Child day-care activities 8891 | | 25658 | 1824(1667;1995) |
| Other retail sale of food in specialised stores 4729 | | 3305 | 1785(1385;2297) |
| Fitness facilities 9313 | | 3741 | 1764(1388;2239) |
| Residential care activities for the elderly and disabled 8730 | | 66609 | 1746(1649;1848) |
| Other credit granting 6492 | | 3230 | 1734(1337;2247) |
| Manufacture of air and spacecraft  3030  and related machinery | | 5251 | 1714(1396;2103) |
| Public order and safety activities 8424 | | 53021 | 1688(1582;1801) |
| **All sectors** | | **4390832** | **1178(1168;1188)** |
| **General population** | |  | **816** |

Table 5: 14-Day incidence of COVID-19 in sectors with the highest incidence at Level 5 Period 4

| Period 4: 29 September–12 October 2020 | | |  |
| --- | --- | --- | --- |
| DESCRIPTION | NACE Code | Employees | Incidence (95%CI) |
| Activities of football clubs | 93121 | 3605 | 1609(1246;2076) |
| Activities of leagues and sports federations | 93191 | 1961 | 1428(988;2060) |
| Other human resources provision | 78300 | 4320 | 1250(959;1629) |
| Fitness facilities | 93130 | 3707 | 1187(884;1591) |
| Other retail trade of food in specialised stores | 47299 | 2857 | 1155(822;1620) |
| General ordinary secondary education | 85311 | 159055 | 1143(1092;1196) |
| General construction of office buildings | 41202 | 1756 | 1139(736;1759) |
| General social services with accommodation | 87902 | 2500 | 1080(742;1570) |
| Production of shows by artistic ensembles | 90012 | 4924 | 1056(806;1383) |
| Other cleaning activities | 81290 | 2846 | 1019(709;1463) |
| **All sectors** |  | **4390750** | **573(566;580)** |
| **General population** |  |  | **423** |
| Period 5: 6–19 October 2020 | |  |  |
| DESCRIPTION | NACE Code | Employees | Incidence (95%CI) |
| Manufacture of weapons and ammunition | 25400 | 2183 | 2657(2060;3422) |
| General construction of office buildings | 41202 | 1761 | 2556(1914;3406) |
| General ordinary secondary education | 85311 | 159806 | 2478(2403;2555) |
| Other human resources provision | 78300 | 4326 | 2381(1967;2880) |
| Activities of football clubs | 93121 | 3556 | 2250(1811;2793) |
| Activities of leagues and sports federations | 93191 | 1956 | 2250(1678;3010) |
| Distribution of gaseous fuels through mains | 35220 | 1881 | 2126(1563;2886) |
| Activities of medical laboratories | 86901 | 5159 | 1977(1631;2395) |
| Nurseries and day-care centres | 88911 | 22564 | 1950(1778;2139) |
| Motion picture projection activities | 59140 | 1633 | 1837(1287;2615) |
| **All sectors** |  | **4390832** | **1178(1168;1188)** |
| **General population** |  | | **816** |

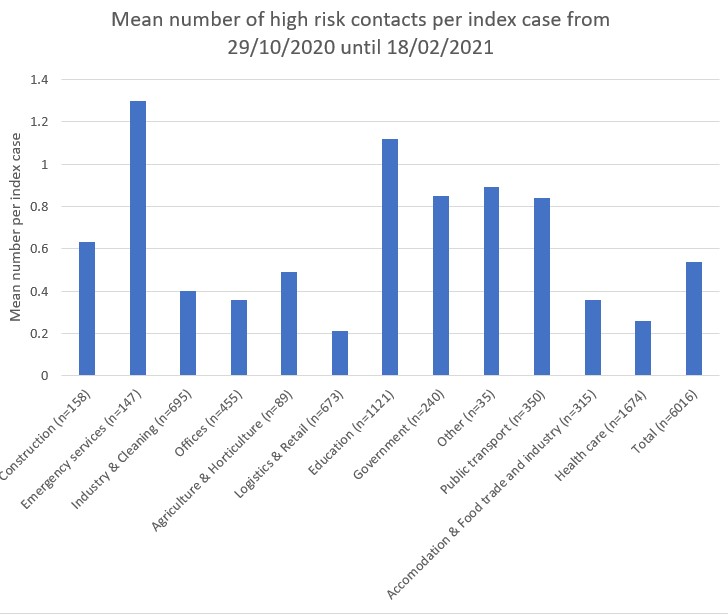

Figure 6: The mean number of high-risk contacts per index case by segments under surveillance

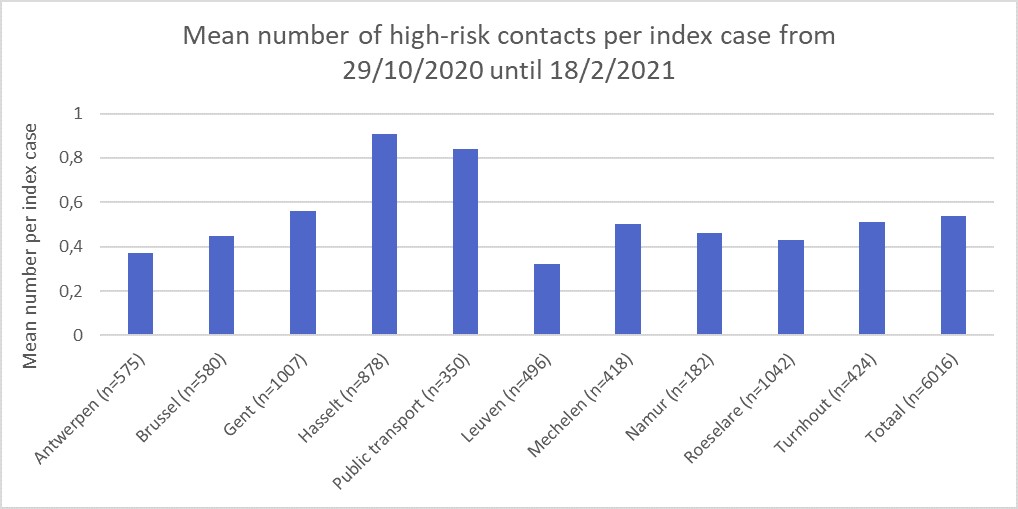
 Figure 7: The mean number of high-risk contacts per index case by region
